# Reducing stillbirth in high burden settings using biomarkers and ultrasound technologies: protocol for the multi-centre prospective iTECH cohort study

**DOI:** 10.64898/2026.06.22.26356223

**Authors:** Sam Ali, Winfred Nakato, Josephine Tumuhamye, Sheila Nabweyambo, Obondo J. Sande, Rose Mukisa-Bisoborwa, Wessel Ganzevoort, Sanne J. Gordijn, Marcus J. Rijken, Kerstin Klipstein-Grobusch, Josaphat Byamugisha, Aris T. Papageorghiou

**Author notes:** Corresponding author: Dr. Sam Ali, Department of Obstetrics and Gynecology, Makerere University Hospital, Makerere University, P.O.Box 7062, Bativa Road, Kampala, Uganda.

## Abstract

**Introduction:** Stillbirth prevention requires reliable detection of potential causes for timely interventions. Currently, there is no effective screening strategy to identify fetuses at risk of stillbirth. Prognostic models have been proposed as a potential solution, but there is a shortage of robust, clinically applicable models in low- and middle-income countries. Early birth is frequently initiated without proper risk stratification, leading to increased neonatal and infant morbidity and mortality. This study aims to develop and validate multi-modal multivariable prediction models for stillbirth and pathologies that lead to stillbirth (preeclampsia & fetal growth restriction) using widely accessible and cost-effective markers. Stakeholder perspectives will also be assessed.

**Methods and analysis:** This multi-center prospective cohort study is running in four high volume regional referral hospitals in Uganda: Kawempe, Hoima, Lira, and Mbale. We will enroll at least 6,075 pregnant women attending routine antenatal care (ANC), above 13 years of age, and ≥11 weeks of gestation. Data and biological samples will be collected at 11-23 weeks, 35-37 weeks and at birth in all women. In a subset of women, additional measurements will be obtained between 24-34 weeks and 38-42 weeks to allow for spread of the data across the full spectrum of pregnancy. This data will enable us to investigate the physiological changes with gestational development in healthy or unhealthy pregnancies, to guide future monitoring and management of women and establishment of reference values for novel markers. The placenta will be collected for histopathological analysis in women diagnosed with intrauterine fetal demise at ≥20 weeks of gestation, stillbirth nearmiss and their corresponding controls.

Data on socio-demographics, obstetric history, current pregnancy conditions, and tests such as maternal hemodynamics, ultrasound, and biochemical markers will be collected from each participant, and used to develop regression and machine learning prediction models. Models will be validated and evaluated by comparing their calibration plots, precision and recall, F1 scores and accuracy, aiming for less complexity and reliable predictions. Emerging models will be translated into software as a medical device (SAMD), while taking into account user experiences, regulatory requirements, data pipelines in clinical workflows and user-friendly interfaces that facilitate access and the interpretation of outputs, to allow for seamless integration into existing electronic health information systems and decision support tools.

To assess stakeholder perceptions, we will employ an exploratory qualitative component using focus group discussions, semi-structured and key informant interviews. The sample will include 81 purposively selected women and their partners who use maternity care services, local leaders and healthcare providers in and out of the four hospitals implementing iTECH in Uganda. Qualitative data will be audio recorded, transcribed verbatim and thematic analysis performed using Nvivo 12.

**Ethics and dissemination:** The study protocol was approved by Makerere University School of Medicine Research Ethics Committee (Mak-SOMREC-2022-535) on March 1, 2023 and the Uganda National Council for Science and Technology (HS2762ES) on April 6, 2023. We will disseminate the findings through scientific publications, stakeholder engagements, Wellcome Leap program meetings, professional bodies, reproductive maternal newborn child and adolescent health coalition platform, Ministry of Health Uganda, district and hospital meetings, webinars, and conferences.

**Strengths and limitations of this study:**

- large sample of women recruited from a general obstetric population to inform stillbirth and related outcomes prediction models based on multi-modality data.
- measurements taken to help understand physiological changes with gestational development to guide future monitoring and management of pregnant women.
- histopathology performed in selected cases and controls to contribute to the understanding of mechanisms that lead to stillbirth and refine model development.
- of active tracking mechanisms like phone calls, short message texts and locator forms to remind mothers of their scheduled appointments and mitigate participant drop-offs.
- care for women included in this study may differ from that received by other women attending routine ANC: this may lead to lower adverse pregnancy outcome rates in this cohort than usual in the setting.

## INTRODUCTION

Up to two million babies are stillborn annually in late gestation (≥28 weeks) worldwide. This is thought to be an underestimate of the true burden given variations in the definitions of stillbirth across settings and poor reporting in many countries [1,2]. While high-income countries have registered significant reductions in stillbirth rates in the past two decades, the narrative is different for sub-Saharan Africa: stillbirth rates have instead increased. For example, in Uganda, there was an 8.6% increment between 2000 and 2019 [2].

The causes of stillbirths are heterogeneous- congenital anomalies and placental dysfunction being key contributing factors [3,4]. Significant decreases in stillbirths will require a well-rounded solution to all known preventable causes. Globally, 7.4% of stillbirths are attributed to congenital abnormalities, though regional disparities exist [5]. In the United States, birth defects account for over 14% of stillbirths [3]. Anomalous fetuses have a significantly elevated stillbirth risk (55 per 1,000 compared to 4 per 1,000 for non-anomalous fetuses; (aOR 15.17, 95% CI: 11.03-20.86)) [6]. The largest proportion of stillbirths, caused by congenital anomalies, can be managed and prevented by early detection [7,8].

Among small non-anomalous fetuses, the common underlying mechanism leading to stillbirths is placental dysfunction attributed to various placental pathologies, maternal vascular malperfusion being the most common and understood lesion. The underlying pathophysiological mechanisms that lead to stillbirths in normal-sized babies are less well understood and a high proportion are classified as unexplained [4]. Existing evidence indicates that placental dysfunction may also have a vital role in this subgroup [9–11]. Related to this, biomarkers of placental function such as human growth hormone (hGH)/chronic somatomammotropin hormone (CSH) genes and placental proteins; soluble fms-like tyrosine kinase 1/placental growth factor (sFlt-1, PlGF) have shown promise [12–14]. For example, growth-restricted babies have low levels of hGH/CSH gene expression [12].

During pregnancy, the maternal cardiovascular system undergoes structural and functional modifications to provide adequate uteroplacental perfusion and nutrition to facilitate normal fetal growth and development [15]. Maternal cardiac adaptation is correlated with physiological and morphological adaptation in the placenta. For instance, uterine spiral artery remodeling physiologically reduces maternal systemic vascular resistance [10]. First-trimester studies have demonstrated that abnormal maternal hemodynamic profiles indicate increased risks of clinical manifestations of fetal growth restriction (FGR) and preeclampsia (PE). Increasing evidence also suggests that underlying maternal cardiac dysfunction is associated with placental dysfunction. Therefore, further exploration of maternal hemodynamics in all trimesters could provide insights into the mechanisms that lead to stillbirth, thus providing an opportunity to improve management outcomes [16–19].

Preventing stillbirth requires timely identification of risk factors to enable effective interventions. At present, no effective screening strategy to detect deprived fetuses at risk of stillbirth exists. Prognostic models to more precisely estimate a woman’s individual risk of stillbirth have been proposed as a potential solution, but there is a shortage of robust, clinically applicable models in low-resource settings [20]. Iatrogenic early birth is often initiated in fear of the worst, however without proper risk stratification resulting in a shift towards neonatal and infant morbidity and mortality. To date, potential markers of clinical importance to predict and prevent stillbirths in low- and middle-income countries (LMICs) have not been well studied.

We have undertaken several studies to address this gap: first, our review of available literature on the prognostic accuracy of Doppler ultrasound for adverse perinatal outcomes revealed a lack of evidence to guide how Doppler ultrasound should be used in LMICs [21]. The review informed a large prospective cohort study which showed that low cerebroplacental ratio (CPR) was strongly associated with stillbirth (OR 4.82, 95% CI 1.09–21.30). CPR and middle cerebral artery (MCA) PI <5^th^ percentile were independently associated with adverse perinatal outcomes [22,23]. A model combining maternal characteristics with MCA PI or CPR had AUCs of 0.78 (95% CI: 0.67 - 0.87) and 0.78 (95% CI: 0.65 - 0.87), respectively, for predicting perinatal death in the development set [24]. The bootstrap corrected AUC was 0.71 with a slope of 0.70 [24], only moderate and below clinically relevant threshold, but this was based on a small sample with low number of events.

Existing models have not been validated in low-resource settings and have low predictive performance [20,25,26]. Most include maternal and ultrasound features, whereas those including biochemical markers have the highest predictive performance [20,25,26]. Large scale multi-modal stillbirth prediction studies are required. Such complex models may benefit from machine learning modeling techniques given their ability to handle a large number of datasets and high-dimensional data, and complex interactions and none-linear relationships between variables [27].

Therefore, the iTECH project aims to use traditional statistical and machine learning methods to develop and validate prediction models for stillbirth and pathologies that lead the stillbirth (PE and FGR) based on widely accessible and cost-effective markers. Previously, we found that spousal involvement may promote acceptance and use of advanced technologies like Doppler ultrasound, but healthcare providers required more capacity building on how to use it for the benefit of mothers [28]. Stakeholder engagements will continue as an important component of the iTECH project.

## METHODS AND ANALYSIS

### Study design

This is a multi-center prospective observational cohort study. A qualitative component will be embedded to gain stakeholder insights on feasibility, acceptability and sustainability of the novel technologies being tested, perceptions and experiences of stillbirth, and their views on how the current ANC model could be transformed to deliver healthcare services efficiently.

### Study Setting

Participants will be recruited from four Hospitals in Uganda: Kawempe National Referral Hospital (NRH), Hoima Regional Referral Hospital (RRH), Lira RRH, and Mbale RRH. Kawempe NRH (about 21000 births annually) is a 200-bed women’s and children’s hospital in Kampala, the capital city of Uganda, but serves a heterogenous population consisting of the urban poor and those with average income. Hoima RRH (bed capacity= 300, total annual deliveries= 7,678 births in 2020/2021) is 230 km west of Kampala, and serves over three million people in the Greater Bunyoro Region. Lira RRH (annual outpatient attendance of 100,000 patients and approximately 6,000 births annually) is 340 km from Kampala and a referral center for the major districts in Northern Uganda. Mbale RRH (52 maternity beds and about 8000 births annually) is 214 km to the east of Kampala, serving the districts in the Mount Elgon zone. These four hospitals are comparable in regards to population risk profiles, infrastructure, and services offered.

According to the Ministry of Health Annual Health Sector Report 2023/2024 and District Health Information System (DHIS2) report, the selected regions had the highest burden of stillbirth and perinatal mortality rates (PMR) in Uganda [29]. Bunyoro and Kampala had disproportionately higher PMR fresh stillbirth (FSB) as the biggest contributor. The PMRs of Lango (northern) region, Bugisu (eastern) region, Bunyoro (western) region and Kampala (central) were 12.7/1000 births, 11.6/1000 births, 22.7/1000 births, and 35.8/1000 births, respectively. The national stillbirth rate stands at 11.9 per 1000 livebirths. These sites were selected based on the national vital statistics on perinatal health, and existing infrastructure and expertise to conduct the study. The strategic selection of the recruitment sites secures both regional representation in the country, and the robustness and feasibility of this study because the sites are high-volume health facilities serving disparate populations.

### Participants

Eligible participants will be pregnant women ≥11 weeks of gestation, regardless of their risk profiles for adverse pregnancy and neonatal outcomes, attending routine ANC at the recruitment sites, above 13 years old, must have conceived naturally, provided written informed consent to participate in the study, and should be planning to deliver from the study site. In addition, emancipated minors (individuals below the age of majority who are pregnant, married, have a child or cater for their own livelihood) have been included in this study given the significant proportion of teenage pregnancies in the country, and the need for an inclusive research and clinical tools that accommodate all pregnancy populations that attend ANC in Uganda. According to the United Nations, there is need to ensure universal access to sexual and reproductive health services among adolescents and track their birth rates as an important indicator for sustainable development goals (SDGs) target 3.7 [30]. The TRIPOD + AI statement also emphasizes that models should be developed in a way that do not adversely discriminate against individuals or groups of individuals based on attributes such as age, race/ethnicity, sex/gender, or socioeconomic status, and does not create inequalities in healthcare provision [27].

Gestational age estimate will be based on an early ultrasound using crown rump length (CRL) measurements at 11^+0^ to 13^+6^ weeks of gestation or a late dating scan using measurements of the head circumference (HC) and femur length (FL) at 14^+0^ to 23^+6^ weeks [31–33]. Pregnant women with major fetal anomalies at ultrasound, multiple pregnancies, pregnancies with complications (fetal miscarriage, embryonic miscarriage, anembryonic miscarriage, ectopic pregnancies, among others), with a gestational age of ≤10 weeks at ultrasound scan and those participating in other clinical trials or studies where interventions used might interfere with the iTECH study procedures or analysis will be excluded. Fetuses with major fetal anomalies have extremely elevated risk for poor birth outcomes, including stillbirth [6], and will thus be excluded from our study population to minimize bias in our predictions.

### Data Collection

#### Recruitment Process

Potential participants will be approached at ANC clinics and informed about the iTECH study. Participants who provide verbal consent will be screened based on the first set of eligibility criteria. The screening questions will include mother’s age, willingness of the woman to give birth from the study site and type of conception. All potentially eligible women will further be asked to provide written informed consent, and assigned a unique participant identification number. The women will then undergo a detailed ultrasound examination to confirm eligibility. All participants confirmed at ultrasound will proceed with all baseline procedures including direct interviews, maternal hemodynamics assessments and blood draw for routine clinical and research tests as shown in **Figure 1**. Those excluded from the study based on ultrasound findings will be advised to join routine healthcare in the hospital.

**Figure 1.**
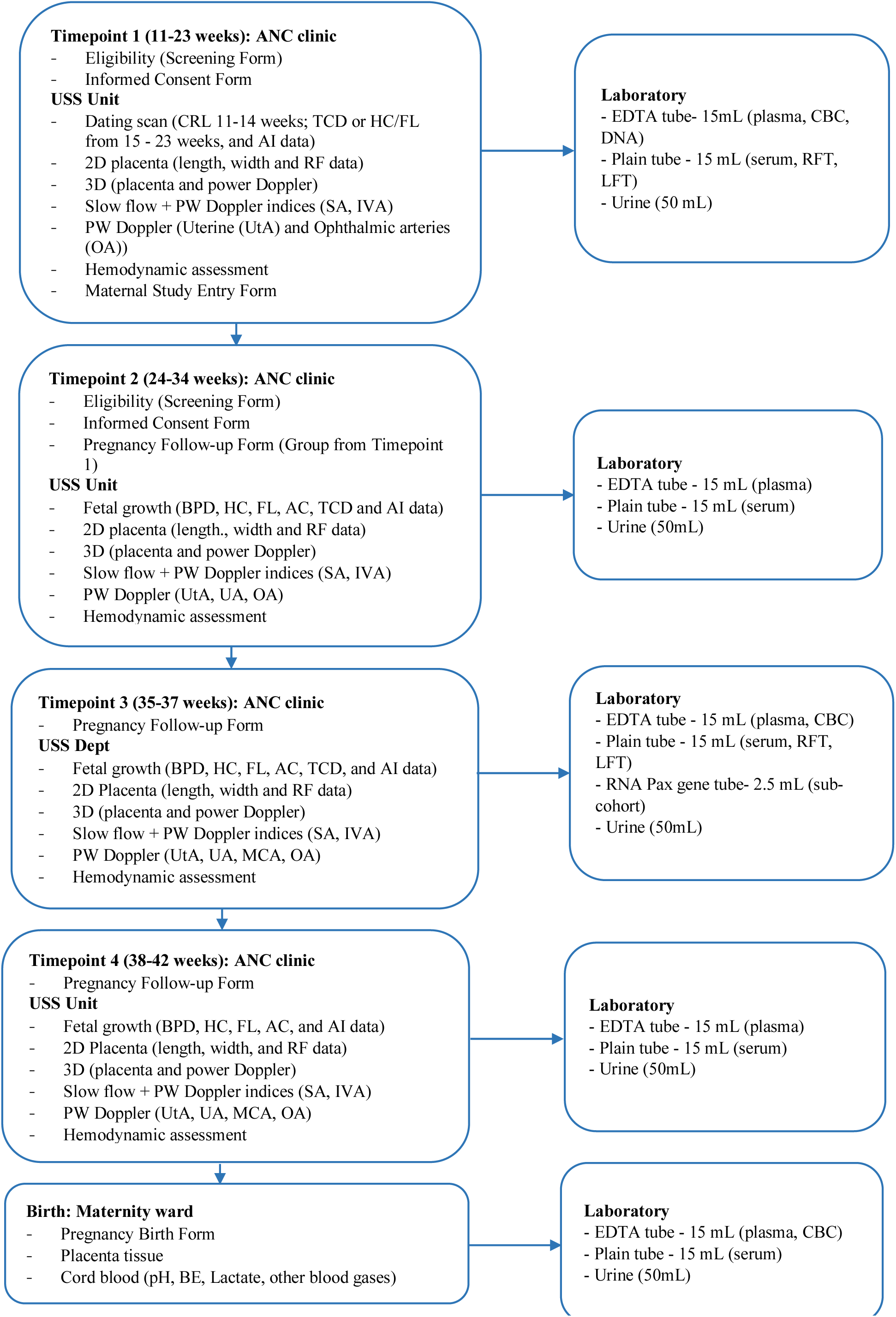
Measurements and biological samples obtained at study recruitment, each follow-up visit and birth

#### Participant Interviews and Follow-up

Once a woman is recruited into the study, she will be interviewed at all study visits. They will be asked about their sociodemographic information, obstetric history, current pregnancy conditions and nutritional status. Privacy and confidentiality will be ensured while conducting the interviews to avoid socially desirable responses. All participants will be longitudinally followed up from the point of recruitment (early pregnancy) through birth, until six weeks after birth. All women will participate at 11-23 weeks and 35-37 weeks. In a subset of women, additional measurements will be obtained between 24-34 weeks and 38-42 weeks to allow for spread of the data throughout gestation. This will allow us to understand physiological changes with gestational development in healthy or unhealthy pregnancies, to guide future monitoring and management of pregnant women and establishment of reference values for novel markers. Women will be randomly assigned to the sub-cohort (subset) using computer-generated random allocation sequence within the Medscinet database as shown in **Table 1**.

**Table 1:** Recruitment and follow-up schedule for participants included in the sub-cohort.

| **Sub-Cohort** | **Visit 1** | **Visit 2** | **Visit 3** | **Visit 4** |
| --- | --- | --- | --- | --- |
| 1 | 11 | 21 | 31 | 36 |
| 2 | 12 | 22 | 32 | 36 |
| 3 | 13 | 23 | 33 | 37 |
| 4 | 14 | 24 | 34 | 37 |
| 5 | 15 | 25 | 35 | 41 |
| 6 | 16 | 26 | 35 | 41 |
| 7 | 17 | 27 | 36 | 40 |
| 8 | 18 | 28 | 36 | 39 |
| 9 | 19 | 29 | 36 | 38 |
| 10 | 20 | 30 | 36 | 38 |

Participants will be given ±1week window period in their appointment dates. A research assistant will contact a participant a week before her actual appointment date to remind her to come back for her scheduled visit. For those that will miss their appointment date and the study visit window has closed, they will be informed to return on their next scheduled appointment date and the missed visit is scored as such. For participants still within their visit window but have failed to be reached by phone, community engagement officers will conduct home visits to remind participants of upcoming study appointments. No study procedures will be conducted outside the individual study window. We will capture pregnancy outcomes at the time of birth, and follow-up all women for neonatal death (within 28 days), maternal death and maternal near miss within 42 days after birth. Women in whom birth outcomes are unknown 6 weeks after their expected due date (EDD) will be classified as lost to follow-up, and this will be minimized by active tracking mechanisms like phone calls, SMS, and conducting home visits using locator forms to remind mothers of their scheduled appointments. All participants will receive contemporary prenatal and postnatal care according to the Uganda National Clinical Standards [34].

#### Ultrasound Examinations

At baseline, fetal biometry measurements including HC, FL, abdominal circumference (AC), and biparietal diameter (BPD) to determine gestational age and fetal anatomy assessment to determine any anomalies will be performed. Doppler examinations (uterine artery (UtA), umbilical artery (UA) and middle cerebral artery (MCA)) and ophthalmic artery (OA) will be done during the study visits according to published standards [23,35]. New measures such as spiral arteries, placental arterioles, quantitative ultrasound (QUS) data and an artificial intelligence algorithm for gestational age determination will be tested. We will examine the placenta using B-mode for placental structure, density, lesion and presence of calcifications, and novel technologies (low velocity flow, 3D volume acquisition) to assess flow and placenta size, to better understand the origin of placental diseases/syndromes and pregnancy complications. Fetal growth assessment with routine biometric measurements will be performed at all follow-up visits. For all examinations, a short video clip (up to 15 seconds) leading to each measurement will be stored, as well as the final still images. To ensure standardization across the four sites, all the sonographers will be uniformly trained and certified, as per our published standards [23].

#### Maternal Hemodynamics Examinations

Maternal hemodynamic examinations will be performed by a well-trained nurse or midwife using the ultrasonic cardiac output monitor (USCOM) and Arteriograph device. The attending staff will explain the procedure to the participant and take preliminary measurements: systolic and diastolic blood pressure using a blood pressure machine (sphygmomanometer), and height and weight using a calibrated weighing scale (SECA). These measurements will be fed into USCOM and Arteriograph machines for computation of cardiac function parameters. USCOM, a non-invasive device utilizing continuous-wave Doppler technology, measures transaortic blood flow to determine parameters such as cardiac output, systemic vascular resistance, and stroke volume. After the participant has been assessed to be calm and relaxed, the study staff will ask her to lie in the left lateral position and the nurse will gently place the probe at the suprasternal notch to measure the transaortic blood flow of the left side of the heart. Arteriograph is a non-invasive cuff device that uses oscillometric technique to measure arterial stiffness. For this examination, the participant will be asked to lie in the supine or fowler’s position at 45 degrees angle to obtain additional hemodynamic parameters like aortic pulse wave velocity, augmentation index and central systolic blood pressure. The study nurses and midwives will be trained and certified to perform USCOM and Arteriograph procedures as per our established standards [36].

#### Biological Sample Collection

Venous whole blood will be collected aseptically from participants and swiveled by gently inverting the bottle 8-10 times immediately after collection to activate the anticoagulant or clotting factor in the tubes to mix with the blood. We will collect 30 mL of blood into 4mL and 10mL EDTA tubes, and into 4mL and 8.5mL SST tubes at every study visit. Additionally, 2.5 mL of blood will be collected using PAX gene blood RNA tubes in a sub-set of women at the 35-37 weeks’ time point. Mid-stream urine (50 mL) will be collected in a sterile urine container at every study visit. At birth, arterial cord blood will be collected into heparinized syringes or tubes for cord blood gas and electrolyte analysis within 30 minutes after birth, using the iSTAT device. In addition, we will collect placentae of stillborn or stillbirth-nearmiss babies, and their corresponding controls. All placentas will immediately be fixed in 10% formalin and transported and later date to the pathology department for analysis.

#### Laboratory Procedures

Blood in 4mL EDTA tube will be used for routine clinical tests; malaria, CBC and HIV tests and the remaining blood stored for future genetic studies, while blood in 10mL EDTA tubes will be used for biomarker analysis. The EDTA tubes will be centrifuged at 2000G (rcf) for 15 minutes at room temperature to separate the plasma which will be aliquoted into six 1mL cryovials. Blood in 4mL SST tubes will be used for routine clinical tests: Hepatitis B test, liver function tests and renal function tests while blood in 8.5mL SST will be used for biomarker analysis. Blood in SST tubes will be centrifuged at 3000 rpm, at room temperature for 10 minutes to separate the serum and aliquoted into six 1mL cryovials each. We will process blood for biomarker analysis within 30 minutes after collection and temporarily store it at −15^0^C to −20^0^C at the sites, before transportation to Makerere University College of Health Sciences Biorepository for long term storage at −80^0^C. Samples will be transported at −20^0^C in leak-proof cooler containers with data loggers to monitor temperature excursions. Liquid biomarker analysis in serum or plasma will be performed at Makerere University Hospital using the Olink Platform (a Proximity Extension Assay (PEA) based technology), to quantify the protein levels. The study lab team will be comprehensively trained and certified in sample collection, labelling, and processing and analysis. All tests will be performed according to the manufacturer’s instructions and study standard operating procedures, including the quality control and assurance manual.

#### Autopsy

A team of pathologists at Mulago NRH, Kampala, Uganda and a perinatal pathologist in The Netherlands will process and investigate the placenta according to the Sampling and Definitions of Placental Lesions: Amsterdam Placental Workshop Group Consensus Statement [37]. Quality checks will be performed by randomly reviewing 10% of cases by a perinatal pathologist. Verbal autopsy will be conducted within six weeks of the event, for participants who experience an early fetal demise (intrauterine death at ≥20 weeks and <28 weeks), stillbirth (intrauterine death at ≥28 weeks) and a neonatal death (0-6 days after birth). The verbal autopsies will be reviewed by a panel of expert specialists consisting of obstetricians, pediatricians/neonatologist to investigate the probable cause of death.

#### Qualitative data collection

Data will be collected through semi-structured individual interviews, focus group discussions, and key informants’ interviews. Individual interviews and focus group discussions are appropriate data collection methods to answer the study research questions aimed at exploring the views and perspectives of key stakeholders.

#### Patient and Public Involvement

Participants were not involved in conceiving this research. During the implementation phase, we will leverage the existing partnerships and alliance membership, reproductive health networks and the power to convene actors, at national and sub-national levels and directly reach community stakeholders including women and girls, influencers, opinion leaders, healthcare providers, and meaningful engagement of males and spouses in the communities - ‘leaving no one behind.’ Mutual discussion and feedback between women, community, and healthcare providers, will culminate in action planning and commitments to address barriers to uptake of ANC.

### Study Variables

The primary outcome is stillbirth defined as fetal death at ≥28 weeks of gestation. Secondary outcomes are predefined as: perinatal death defined as the occurrence of either stillbirth or neonatal death within 6 days of the postnatal period; stillbirth near-misses defined as FGR (birthweight <3^rd^ centile, absent or reversed end diastolic flow in umbilical artery, and birthweight <10^th^ centile with abnormal Dopplers detected antenatally), signs of perinatal hypoxia (Apgar <7 at 5 minutes; umbilical artery PH <7.1, neonatal cardiac resuscitation, neonatal encephalopathy), pre-eclampsia with severe features; neonatal death (death of a baby between day 0 and day 6 of life), placental abruption (clinical diagnosis documented in healthcare records), severe placental lesions (documented in clinical pathology report as high grade or severe maternal vascular malperfusion (MVM) AND/OR high grade or severe fetal vascular malperfusion (FVM)); small for gestational age (SGA) defined as birthweight <10^th^ percentile using International Fetal and Newborn Growth Consortium for the 21st Century (INTERGROWTH-21st) newborn birthweight standards [38]; and fetal growth restriction defined as per the Delphic consensus [39].

Pre-eclampsia defined as per the ISSHP criteria, as elaborated: (blood pressure ≥140/90 mmHg arising de novo after 20 weeks’ gestation accompanied by proteinuria and/or evidence of maternal acute kidney injury, liver dysfunction, neurological features, hemolysis or thrombocytopenia, and/or fetal growth restriction) with repeated episodes of severe hypertension (>160/110mmHg) despite maintenance treatment with multiple antihypertensive agents and/or progressive thrombocytopenia and/or progressively abnormal renal or liver enzyme tests and/or pulmonary oedema and/or abnormal neurological features such as severe intractable headache, repeated visual scotomata, or convulsions and/or non-reassuring fetal status and/or diagnosis of eclampsia; or pre-eclampsia arising at less than 34 weeks of gestation (diagnosis between 20^+0^ weeks gestation and 33^+6^ weeks gestation) [40].

Predictor variables will include data on socio-demographics, obstetric history, current pregnancy conditions, and measurements such as maternal hemodynamics, ultrasound, and liquid biomarkers. Definitions for healthy pregnant women and socioeconomic constraints of fetal and newborn growth (**Table S1** and **Table S2**) will be locally adapted, following validated standards [41–43]. Socio-economic factors will include: education, occupation, employment status, housing tenure and markers of household income (wealth index). The wealth index will be determined by data reduction technique using principal component analysis of a selected set of variables in **Table S2**.

Household wealth index will be summarized into quintiles with the first quintile showing the poorest households and the fifth quintile, the most well-off households.

### Sample size calculation

We estimated the required sample for iTECH study using R package, “pmsampsize”, assuming a stillbirth rate of 17.8 per 1000 live births in Uganda [2] and a discriminative ability of ≥0.9. We specified a model for a binary outcome fitting 30 candidate predictor parameters using a shrinkage factor of 0.9 and ±0.05 precise estimate of the average outcome risk in the population, resulting in 5,062 participants. Given the longitudinal nature of the study, including multiple timepoints for data capture, we inflated the sample by 20% to cater for non-response and any loss to follow-up, giving us up to 6,075 participants needed for enrollment. We aim to include a subset of ≥1,500 participants with additional measurements outside the two primary study windows of 11-23 and 35-37 weeks. For the qualitative work, we will include 81 participants, including 32 women of different categories, 14 male partners, 10 community leaders and 25 healthcare providers. A sample of 81 is estimated to be sufficient to reach data and thematic saturation based on the number of stakeholders required to participate in the study.

### Data management and statistical analysis

Before analysis, variable transformations and imputation for missing data will be done [44]. We will use statistical softwares (R and Python) for analysis. Linear relationships between continuous predictor variables and the outcomes will be checked using fractional polynomials and restricted cubic spline plots, before developing prediction models. We will evaluate various models including logistic regression and explore the use of machine learning techniques such as Random Forest, Decision Tree, Stochastic Gradient Boosting, Classification and Regression Trees, Support Vector Machine, Lasso & Ridge Regression, Elastic Net Regression, among others. At model development, we will first fit a baseline model with maternal characteristics alone; then extend it to include other clinical tests as used in clinical practice. Non-contributing predictors will be excluded by methods such as backward stepwise variable selection using the Likelihood ratio tests at a p-value of 0.157 or LASSO regression with L1 regularization to select features with none-zero coefficients [27]. We shall perform data preprocessing using a standardscaler for continuous variables and one-hot encoding for categorical variables with more than two levels. Data imbalance will be taken care of using oversampling and hybrid sampling techniques such as Synthetic Minority Oversampling Technique (SMOTE), SMOTE with nearest neighbors, SMOTE-TomeLink, among others using the imbalanced learn python library. Where applicable, a case cohort design will be employed to minimize data imbalance.

After model development, we will internally validate the reduced models using 200 bootstrap samples randomly drawn with replacement and/or cross validation with 10-folds, StratifiedKFold CV, or using a 0.2-0.3 sample withholding method, averaging the results over a range of bootstrap samples or the number of folds to lower prediction errors. The difference between the training and test data performance will be used to estimate the optimism, reporting the development and the corrected/adjusted performance measures. Machine learning models will be built using the Scikit-Learn python package. The criteria for comparison models will include: discrimination metrics (precision, recall and F1-score to valuate class-wise performance, AUC - ROC curve), calibration (slope and intercept close to 1 and 0, respectively), and calibration plots of actual and predicted probabilities. The most optimal model, with less complexity, less bias (not overfitted), and with the highest predictive performance (discriminative ability and diagnostic accuracy) will then be selected. Results will be reported as per the TRIPOD+AI statement: updated guidance for reporting clinical prediction models that use regression or machine learning methods [27]. Qualitative data will be audio recorded, transcribed verbatim and coded in two cycles using NVivo 12 to generate new themes and insights using both inductive and deductive approaches. In order to ensure clinical usability of our models, we shall put only clinically relevant features into the model so as to align with standard clinical workflows. Clinicians will also be involved early in the design and testing phases to ensure the model’s outputs (e.g., risk scores, classifications) are presented in a clear and meaningful way that supports decision-making. The final tool will be integrated into existing clinical systems or mobile applications in a way that requires minimal training and does not disrupt existing workflows.

### Ethics And Dissemination

This three-year project will run from October 1, 2022 to September 30, 2025. This protocol has been approved by Makerere University School of Medicine IRB (SOMREC) Ref. Mak-SOMREC-2022-535 on March 01, 2023 and the Uganda National Council for Science and Technology (UNCST) Ref. HS2762ES on April 06 2023. All participants will provide written informed consent before enrolment into the study. Permission was obtained from SOMREC to enroll emancipated minors, and separate consents are being secured for this purpose. The regulatory guidelines in the country regard pregnant teenagers as capable of making independent decisions. However, we anticipate that most of the pregnant teenagers will be staying with their parents and will travel with them to the ANC clinic. Both will be given information about the study as the role of parents will be vital in supporting their children regarding pregnancy and decisions made.

The project data will be electronically captured in the Medscinet (a secure online database), where the data managers directly review all CRFs daily and monthly, for completeness and accuracy. Audit trails will be maintained. Paper based data will be kept under lock and key while electronic data will be password protected with limited access only to authorized study personnel. We will establish a data sharing agreement with our collaborators to ensure restricted data access and regulate the utilization of data generated from this study. We will disseminate the results through scientific publications, stakeholder engagements through In Utero program, professional bodies, reproductive maternal newborn child and adolescent health (RMNCAH) coalition platform, Ministry of Health Uganda technical working groups, district and hospital meetings; media (local media, social media, print media), webinars, conferences, and patient groups.

## DISCUSSION

The stillbirth rates in LMICs have barely dropped in the last two decades. Most stillbirths stem from preventable causes, suggesting that limited rate reductions indicate gaps in existing prevention strategies. At present, a screening strategy for at-risk fetuses recommended by the World Health Organization (WHO) involves a two step-approach of offering an ultrasound to women with a non-reassuring symphysis–fundus height (SFH) measurement to detect growth-restricted babies [45]. The routine screening policy based on SFH (standard of care) is ineffective because it only potentially identifies SGA fetuses, rather than the fetuses at risk of stillbirth [46,47]. SGA fetuses are often undetected by ultrasound, even when detection is prioritized, fetal smallness is not the clear discriminatory parameter of pathology. This is because SGA fetuses can be healthy whilst appropriate for gestational age fetuses can suffer from poor placental function.

Prognostic models to more precisely estimate individual risk for poor pregnancy outcomes in order to facilitate allocation to the appropriate care pathway have been proposed as a potential solution, but there is a shortage of robust clinically applicable models in LMICs [20,25,26]. This project aims to deliver on multi-modal multivariable prediction models for stillbirth and other related adverse outcomes like PET and FGR, incorporating maternal factors, maternal hemodynamics, ultrasound markers and biochemical markers. The final model will be presented as a website tool, mobile app and an offline desk-friendly risk score tool. Emerging models will be translated into software as a medical device (SAMD), taking into consideration user experiences, regulatory requirements, data pipelines in existing clinical workflows, to allow for seamless integration into electronic health information systems and decision support tools. User friendly interfaces to facilitate access and interpretation of the outputs and guidelines for its use will be developed.

Translation of the iTECH models for integration and implementation into care pathways will follow a structured process [48], beginning with stakeholder engagement to ensure their needs are well understood. The stakeholders will include healthcare providers such as midwives, obstetricians, medical officers, policymakers from Ministry of Health’s Digital Health division, and regulatory bodies. Next, we will map the existing clinical care pathways to identify the optimal integration points for the tool. The model outputs will be designed for deployment at various levels of care, including community level and regional or national referral facilities, accounting for differences in infrastructure and services at each level. Before full-scale deployment, the models will undergo further evaluation for impact on pregnancy outcomes and costs. Stakeholder engagements will also lead to deeper understanding of individual and community perceptions of stillbirth by shedding light on the emotional, cultural, and societal factors that shape these perceptions. The findings will be valuable to healthcare professionals, support organizations, policy makers and families dealing with stillbirth, ultimately leading to improved care and support for those affected in Uganda, and similar LMIC settings.

### Strengths and limitations of this study

A major strength of this study is the ability to enroll a large sample of women from a general obstetric population attending routine ANC in an LMIC context and collecting multi-modal data using standardized methodologies from all of them to inform multivariable prediction models of stillbirth and related adverse pregnancy outcomes. The placenta will also be collected in a subset of participants, for histopathological analysis and this will further contribute to the understanding of mechanisms that lead to stillbirth and help refine the prediction models.

This will be one of the first studies in an LMIC to utilize the Olink proteomics platform equipped with a fully automated work flow, that was recently set-up at Makerere University Hospital, to explore protein biomarkers predictive of stillbirth, FGR and PE. This advanced technology utilizes exceptionally small sample volumes (as little as 0.1 microliters (μL)), to analyze specimens with remarkable precision in an efficient and environmentally sustainable way [49]. It is important to note that the Olink laboratory facility at Makerere University is a purely discovery platform, thus the promising markers will need to be translated for integration into routine clinical care pathways. Access to serum or plasma biomarker analysis is still very limited in LMICs due to the lack of laboratory diagnostic infrastructure [50], and if available could be very costly. Ultimately, our goal is to identify markers with potential for deployment at the point of care in a cost-effective way as lab on a chip, making these new tests more accessible in underserved regions.

Another advantage of our study is the use of both traditional statistical and advanced machine learning methods to develop multi-modal prediction models. Machine learning methods offer the capability to handle large numbers of datasets and diverse features, model complex interactions and none-linear relationships between the variables, and deliver prediction models in a faster and easier way by automation of processes, among other benefits [27]. However, due to the flexibility and complexity underpinning some machine learning techniques, sometimes the resulting models are not easy to interpret and even the predictors may be hard to identity [27]. Balancing predictive model complexity and the usability, requirements for integration and interpretation of outputs for clinical utility is very critical.

Lastly, we anticipate some participant loss to follow up due to the longitudinal and complex nature of this study and this will be mitigated using active tracking mechanisms. Also, the sample size was slightly inflated to account for this at design stage. In addition, the care for women included in this study may differ from the care other women receive and may lead to lower adverse outcome rates in our cohort than usual in the setting.

## Supporting information

Table S1: Individual characteristics for participants classified as healthy (low-risk) pregnancies

Table S2: Set of variables for computing household wealth index

## AUTHORS’ CONTRIBUTIONS

SA contributed to the study conception, design and drafted the manuscript. WN, and JT drafted the manuscript. SN contributed to protocol development. OJS, RMB, WG, SG, MJR, KKG, JB and ATP and contributed to the study conception and design, edited and approved the final version. SA is the guarantor

## Data Availability

Not Applicable

## ACKNOWLEDGEMENTS

We thank the iTECH study team, and the health authorities for allowing Kawempe NRH, Hoima, Lira and Mbale RRHs to participate in the study. Fosca Tumushabe provided editorial support. We thank Wellcome Leap for funding this study.

## Online supplementary data legends

**Table S1:** Individual characteristics for participants classified as healthy (low-risk) pregnancies

**Table S2:** Set of variables for computing household wealth index

