## Supplementary material for "Reducing stillbirth in high burden settings using biomarkers and ultrasound technologies: protocol for the multi-centre prospective iTECH cohort study": Table S1: Individual characteristics for participants classified as healthy (low-risk) pregnancies

a) Aged ≥18 and <35 years;
b) BMI ≥18.5 and <30 kg/m2
c) Height ≥ 153 cm;
d) Singleton pregnancy;
e) A known LMP with regular cycles (defined as 28 days ±4 days) without hormonal contraceptive use, or breastfeeding in the 2 months before pregnancy;
f) Natural conception;
g) No relevant past medical history (refer to screening form), with no need for long-term medication (including fertility treatment and over-the-counter medicines, but excluding routine iron, folate, calcium, iodine or multivitamin supplements);
h) No evidence of socio-economic constraints likely to impede fetal growth identified using local definitions of social risk;
i) No use of tobacco or recreational drugs such as cannabis in the 3 months before or after becoming pregnant;
j) No heavy alcohol use (defined as > 5 units (50ml pure alcohol) per week) since becoming pregnant;
k) No more than one miscarriage in the 2 previous consecutive pregnancies;
l) No previous baby delivered pre-term (<37 weeks) or with a birth weight <2500g or >4500g;
m) No previous neonatal or fetal death, previous baby with any congenital malformations, and no evidence in present pregnancy of congenital disease or fetal anomaly;
n) No previous pregnancy affected by pre-eclampsia/eclampsia, HELLP syndrome or a related pregnancy-associated condition;
o) No clinically significant atypical red cell alloantibodies;
p) Negative urinalysis;
q) Systolic blood pressure <140 mmHg and diastolic blood pressure < 90 mmHg;
r) No diagnosis or treatment for anemia during this pregnancy (Hb levels will be monitored throughout pregnancy)
s) No clinical evidence of any other sexually transmitted diseases, including syphilis and clinical Trichomoniasis;
t) Not in an occupation with risk of exposure to chemicals or toxic substances, or very physically demanding activity to be evaluated by local standards. Also, women should not be conducting vigorous or contact sports, as well as scuba diving or similar activities
