## Supplementary material for "Reducing stillbirth in high burden settings using biomarkers and ultrasound technologies: protocol for the multi-centre prospective iTECH cohort study": Table S2: Set of variables for computing household wealth index

| | |
|---|---|
| a) Main wall material of the dwelling<br>1. Burnt bricks<br>2. Cement/concrete blocks<br>3. Mud and pole<br>4. Unburnt bricks/others.<br>b) Main floor material of the dwelling<br>1. Cemented/tiled flooring<br>2. Mud/earth flooring<br>c) Main roofing material of the dwelling<br>1. Iron sheets<br>2. Tiles<br>3. Concrete<br>4. Grass thatch<br>5. Tins/others<br>d) Water supply.<br>1. Tap water<br>2. Bore hole<br>3. Protected well<br>e) Ownership of transport means<br>1. Vehicle<br>2. Motorcycle<br>3. Bicycle<br>4. Boat/canoe | f) Sanitation facility<br>1. Flushing toilet<br>2. Ventilated pit latrine<br>3. No pit latrine<br>g) Energy for cooking<br>1. Electricity-National grid<br>2. Electricity-solar<br>3. Gas<br>4. Paraffin<br>5. Charcoal<br>6. Fire wood<br>h) Electricity for lighting<br>i) Ownership of agricultural land<br>j) Radio<br>k) Television<br>l) Telephone<br>m) Refrigerator<br>n) A bank account.<br>o) Domestic servant<br>p) Average persons per sleeping room |
